# Evaluation of in-house molecular assays for detection of HIV-1 proviral DNA

**DOI:** 10.1101/2025.07.31.25332407

**Authors:** Jaqueline Helena da Silva Santos, Cintia Mayumi Ahagon, Gabriela Bastos Cabral, Giselle Ibette Silva Lopez-Lopes, Luís Fernando de Macedo Brígido

## Abstract

HIV-1 proviral DNA is a critical marker for the early diagnosis of vertical transmission, especially when viral load RNA is suppressed by antiretrovirals. Our objective was to evaluate the performance of a qualitative qPCR to detect HIV-1 proviral DNA. Using the TaqMan system, the assay targets the Integrase (IN), Long Terminal Repeat (LTR) regions of HIV-1, and the CCR5 gene as an internal control. To improve sensitivity, a Nested qPCR step was implemented. ACH-2 cells were used to construct a standard curve and standardize the assays. The assessment of the diagnostic sensitivity and specificity of qPCR was carried out on 125 clinical samples, with the performance of Nested qPCR evaluated on 47/125 clinical samples. qPCR had a detection limit of 40 copies/reaction of HIV-1 proviral DNA, a sensitivity of 85.3%, and a specificity of 100%. Nested qPCR increased sensitivity to 30 copies/reaction. The assays proved to be effective in detecting HIV-1 proviral DNA, identifying a variety of subtypes and CRFs of the virus circulating in Brazil, and applicable to a variety of cases, including those with suppressed plasma viremia.

## 1. Introduction

The early diagnosis of Human immunodeficiency virus (HIV-1) is critical, as it allows the timely initiation of antiretroviral therapy (ARV), which in turn substantially reduces mortality and considerably improves virological control, limiting reservoir size (Violari et al., 2008; Shiau et al., 2017). Given the reported benefits of early initiation of ARVs, early diagnosis of HIV-1 in newborns and timely linkage to treatment are lifesaving interventions. Serological tests do not allow the early diagnosis of HIV-1 infection in children younger than 18 months of age due to the transplacental passage of maternal antibodies (Yilmaz, 2001). WHO recommends the use of molecular tests for the diagnosis of children exposed to HIV-1, quantification of viral RNA - viral load (VL), or detection of HIV-1 proviral DNA, both tests are equally recommended. However, despite the high accuracy of tests that detect HIV-1 RNA, their sensitivity can be affected both by ARV used to prevent mother-to-child transmission or treatment, which reduces HIV-1 RNA levels (Penazzato et al., 2014). Qualitative tests for detecting HIV-1 proviral DNA are less affected and are widely implemented for early childhood diagnosis of HIV-1 in settings with limited resources (UNITAID, 2015).

In addition to early pediatric diagnosis, detection of HIV-1 proviral DNA offers other important applications, such as (1) diagnosis in cases of infection but no RNA detection ( virological controllers); (2) diagnosis of patients with indeterminate serological test results or, (3) cases where HIV-1 RNA is not detectable in plasma due to ARV use for prophylaxis or early treatment and; (4) detection and quantitation of proviral DNA to examine viral reservoirs, as to assess the impact of long-term treatment (Finzi et al., 1997, Chun et al., 1998, Zhao et al., 2002, Gibellini et al., 2004).

Commercially available assays for HIV-1 proviral DNA detection require specific kits and equipment that may be beyond the scope of many resource-limited settings. An in-house Real-time PCR (qPCR) using more widely available and easy-to-perform equipment, requiring less operator training, would provide an attractive alternative to currently available commercial assays (Fiscus et al., 2006).

To support diagnostic expansion, a rapid (<4 h), sensitive, specific, and applicable assay for a variety of HIV-1 subtypes and recombinants was evaluated. The assay for detecting HIV-1 proviral DNA was developed using DNA obtained from peripheral blood mononuclear cells (PBMC) collected directly from the buffy coat, without the need for the meticulous Ficoll-Hypaque step for PBMC separation, as described in several other studies (Désiré et al., 2001; Zhao et al., 2002; Lillo et al., 2004).

Aiming to increase sensitivity, an extra Nested qPCR step was developed and evaluated to combine the accuracy and precision of qPCR and the sensitivity of a Nested PCR, a methodology, used by Pasternak et al., 2008 and Kibirige et al., 2022 in previous works.

## 2. Methodology

### 2.1. Clinical samples

The study included 125 anonymized clinical samples, referred by the services provided by the Retrovirus Laboratory – HIV Genotyping, Virology Center, Institute Adolfo Lutz Central in São Paulo, Brazil. Whole blood collection (EDTA tube) was performed at clinical services and sent to the laboratory. The tube was centrifuged to separate the plasma and the leukocyte buffer; and stored at −70°C until use.

The 125 samples were classified into two groups: Group 1-Samples of patients living with HIV-1: adults with different range VL results (aviremic: VL not detected or < lower limit of detection n = 22; low viremia VL <1000 copies/mL n= 27; high viremia VL > 1000 copies/mL n= 23) and pediatrics (vertical transmission) n= 30. Group 2-Samples from pediatric patients exposed to HIV-1 with discarded diagnosis (two or more tests of VL TND collected in the absence of ARV therapy) n=17 and samples from volunteers known to be HIV-1 negative (4th generation rapid test negative) n = 6.

The patient’s clinical data regarding the history of HIV-1 VL tests and TCD4+ lymphocyte counts were obtained through the Laboratory Test Control System of the National Network for TCD4+/TCD8+ Lymphocyte Counting and HIV Viral Load (SISCEL). Information on the treatment used by the patient was obtained through the Medication Logistic Control System (SICLOM). Both digital platforms, SISCEL and SICLOM, are platforms managed by the Department of Surveillance, Prevention and Control of STIs, HIV/AIDS, and Viral Hepatitis, ensuring the reliability and updating of data.

This study was reviewed and approved by the Institutional Ethical Committee (CEPIAL 71856617.4.0000.0059).

### 2.2. DNA extraction

DNA extraction was performed from PBMC, obtained from the leukocyte buffer, using a commercial extraction kit (QIAamp DNA Mini Kit – QIAGEN, Germany), according to the manufacturer’s recommendations. The extracted DNA was measured in a spectrophotometer (DeNovix DS-11, Uniscience, Brazil) to obtain concentration and purity (OD 260/280).

### 2.3. Amplification and detection: real-time PCR

The qPCR was performed using the TaqMan system, the primers and probes directed the Integrase (IN) and Long Terminal Repeat (LTR) regions, highly conserved regions of the HIV-1 genome, and as endogenous internal control, we used the chemokine receptor 5 gene Human CC (CCR5), a highly conserved gene in all human ethnicities (Malnati et al., 2008; Cillo et al., 2014). The primers and probes (ThermoFisher, USA) were used at a final concentration of 300nM and 200nM, respectively. Their sequences and location in the HIV-1 genome are shown in Table 1.

**Table 1:**
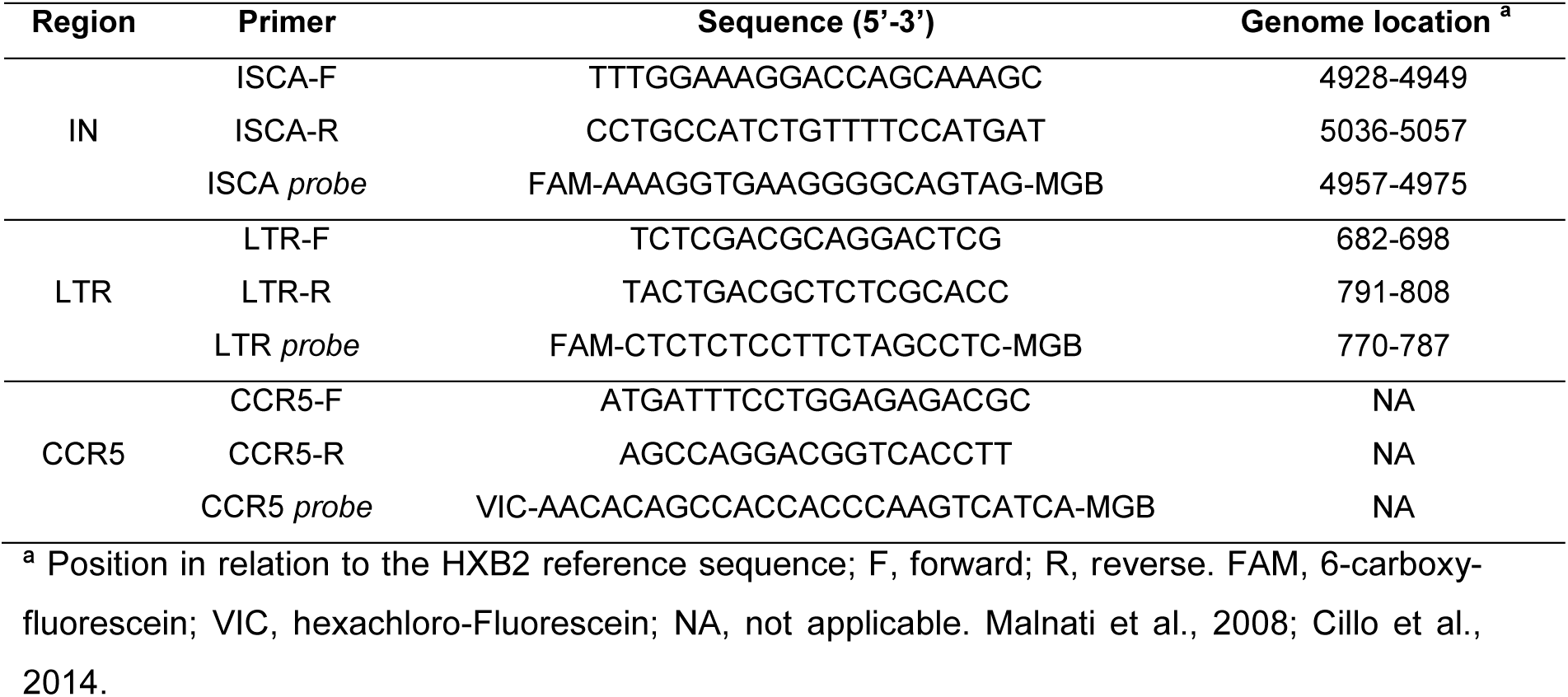
Primers and probes used in real-time PCR with indication of the respective sequence and its location in the HIV-1 genome.

The reaction volume of 20 µL, contained: 10 µL GoTaq ® Probe qPCR master mix with dUTP (Promega, USA), 0.05 µL carboxy-rhodamine (CXR) reference dye, 0.6 µL primer forward and reverse at 10µM, 0.8 µL probe at 5µM, 2.95 µL nuclease-free water and 5 µL extracted DNA. The amplification cycle conditions were as follows: 1 cycle of 94°C for 2 minutes followed by 45 cycles of 94°C for 15 seconds, and 60°C for 1 minute, using the Applied Equipment Biosystems ™ 7500 Real-Time PCR System. Run analysis was performed using the 7500 v.2.3 software (TermoFisher, USA).

Target detection reactions were performed separately. Samples were tested in duplicate for HIV-1 targets. Positivity was determined by observing the exponential curve and the Ct value <40. A sample was considered positive even if it was detectable in only one of the replicates.

### 2.4. Nested chain reaction by conventional polymerase followed by real-time polymerase chain reaction (Nested qPCR)

The first PCR for amplification of the IN region and LTR was performed using standardized laboratory protocols. The reagent volumes were the same for both protocols modifying only the primer pairs (IN primers KVL 70 and KVL 84; LTR primers Seq1f and SK 431). Their sequences and location in the HIV-1 genome are shown in Table 2.

**Table 2:**
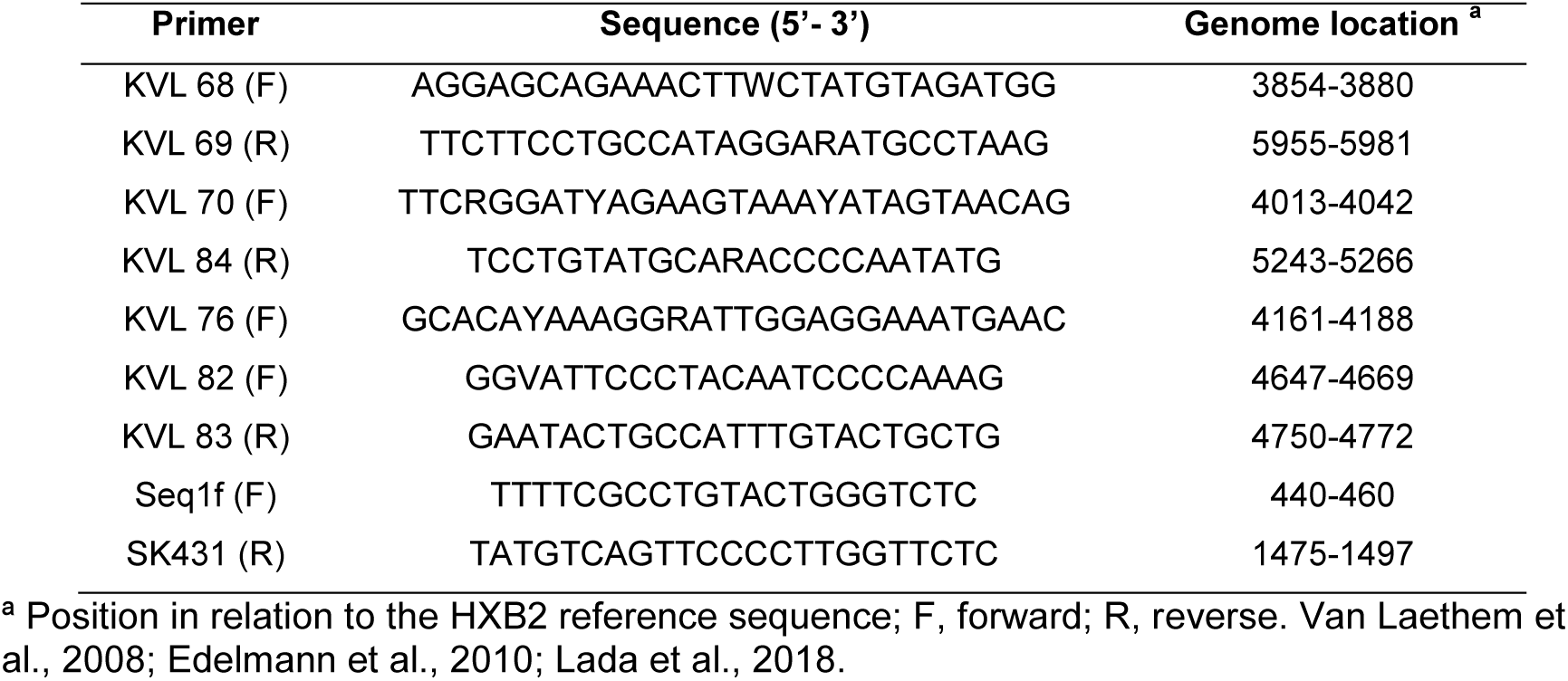
Primers used for conventional PCR amplification and genetic sequencing of the IN and LTR region of HIV-1, indicating the respective sequence and its location in the virus genome.

The first reaction was performed using: 12.5µL master GoTaq mix Colorless 2x (Promega Bioscience, USA), 1µL of 10µM forward and reverse primer, 8µL of nuclease-free water and 2.5µL of extracted DNA. The cycling conditions were: 94°C for 2 minutes, followed by 15 cycles of 94°C for 30 seconds, 53°C for 30 seconds, 72°C for 1 minute and 30 seconds, ending with 1 cycle of 72°C for 7 minutes.

After amplification, a volume of 5µL of the 1st PCR product was used for a second amplification, now by qPCR using the IN, LTR, and CCR5 protocols described previously (item 2.3). The summarized scheme of the Nested qPCR protocol is presented in 1. The protocol summary scheme Nested qPCR is presented in Figure 1.

**Figure 1.**
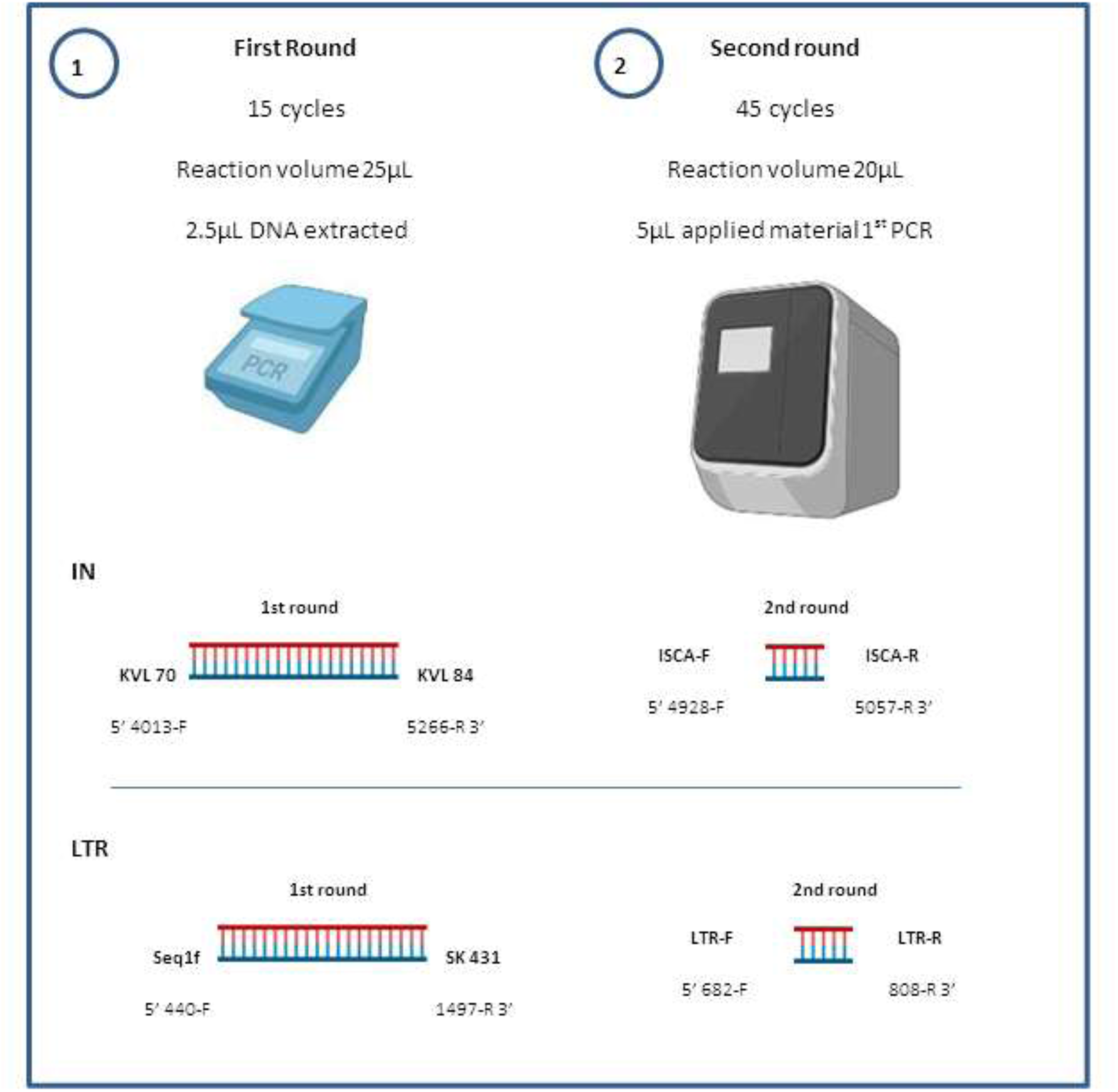
Summary schematic of the Nested qPCR assay. Produced by the author himself using the Biorender.com tool (2022)https://app.biorender.com/biorender.

### 2.5. Standard Curved Construction

Construction of the standard curve was performed from the serial dilution of ACH-2 cells (NIH ARP-349, lot 150053). The ACH-2 cell is a CD4-, CD5+ T-cell clone, with a single copy of HIV-1 proviral DNA integrated per cell. ACH-2 cells were diluted in RPMI 1640 culture medium to obtain a panel of 5 - 500,000 copies.

### 2.6. Detection limit: real-time PCR

We estimated the limit of detection (LOD) using ACH-2 cell dilutions at concentrations of approximately 50, 40, 30, 10, and 5 copies. Assays were performed on 20 replicates of each concentration for each HIV-1 target. The LOD was determined as the lowest detectable concentration in 95% of the replicates tested.

### 2.7. Inter-assay precision and reproduction

Analysis of inter-assay precision and reproduction in samples with different DNA concentrations were performed using dilutions of the ACH-2 cell in the concentrations of 5×10^3^ at 5×10^0^, samples were processed in duplicates, for five days. Since the amount of DNA is inversely proportional to the cycle of amplification at which the target was detected, i.e., the higher the concentration of DNA, the lower the Ct value (Higuchi et al., 1993), the classification of positivity was arbitrarily considered by Ct as: High (< 28); Average (between 30-34); Low (35-37) and Scarce (38 to 39).

### 2.8. Real-time PCR performance

The sensitivity and specificity of the assay were evaluated with 102 samples from HIV-1 infected patients with different levels of plasma viremia and 23 samples from HIV-1 negative individuals, 17 from pediatric patients exposed to HIV-1 but with discarded diagnosis, considering VL results and 6 samples from HIV-1 negative volunteers.

### 2.9. Amplification of HIV-1 proviral DNA by nested PCR and Sanger sequencing

To evaluate the influence of circulating subtypes and recombinants in qPCR amplification efficiency, the entire integrase region (approximately 1106 base pairs) was amplified by nested PCR and sequenced by the Sanger method, according to the protocol developed by Van Laethem, et al 2008. The sequences of primers used in PCRs and sequencing are described in Table 2.

The first reaction was performed using: 12.5µL master GoTaq mix Colorless 2x (Promega Bioscience, USA), 1.5 µL of 10µM forward (KVL 68), and reverse (KVL 69) primer, 7µL of nuclease-free water and 5µL of extracted DNA. The cycling conditions were: 94°C for 3 minutes, followed by 35 cycles of 94°C for 45 seconds, 58°C for 20 seconds, 72°C for 2 minutes and 30 seconds, ending with 1 cycle of 72°C for 7 minutes. For the second amplification, 12.5µL master was used mix of GoTaq Green 2x (Promega Bioscience, USA), 1µL of 10µM forward (KVL 70), and reverse (KVL 84) primers, 8µL of nuclease-free water and 2.5 µL of the material amplified in the first reaction. The 2nd PCR was performed using cycling from 94°C for 2 minutes followed by 35 cycles from 94°C for 15 seconds, to 58°C for 20 seconds, 72°C for 2 minutes, and a final step at 72 °C for 7 minutes. Both amplifications were performed in a thermal cycler. Verity ™ 96-Well Fast Thermal Cycler. The nested PCR product was visualized on a 1% agarose gel in 0.5X TBE buffer (Tris/Borate/EDTA) stained with Sybr Safe® (Life Technologie, USA) using Low DNA Mass Ladder (Life Technologie, USA) as molecular weight marker and standard for concentration.

After the amplification of proviral DNA, the next step was genetic sequencing using the Sanger method. The assay was performed using four primers (KVL 76, KVL 84, KVL 82, and KVL 83) to completely cover the integrase region. A mix was prepared for each primer containing: 4µL of sequencing buffer 2.5X Sequencing Buffer (Applied Biosystems, USA), 0.5µL Big Dye™Terminator cycle sequencing ready Reaction – ABI Prism ® (LifeTechnologies, USA), 3µL primers (1 pm/µL), 1µL nuclease-free water (Invitrogen) and 1.5µL amplified product (10-20ng). The reaction was performed following cycling conditions: 25 cycles of 96°C for 10 seconds, 50°C for 5 seconds, and 60°C for 4 minutes. After the sequencing reaction, the material was purified and precipitated with ethanol and sodium acetate, after precipitation 10µL of formamide was added Hi -Di (Life Technologies, USA), and then the denaturation step was performed using a thermocycler at 95°C for 3 minutes. Sequencing was performed using the ABI 3500XL Genetic automatic analyzer Analyzer (Life Technologies, USA). The chromatograms were manually edited using the *RECall program* (http://pssm.cfenet.ubc.ca).

### 2.10. Statistical analysis

The data obtained were recorded and organized in an Excel spreadsheet (Microsoft, 2010) and analyzed using the STATA program (v8 Stata Corp, USA). Continuous variables are presented as median and interquartile range (IQR: 25-75) and confidence interval (CI) as 95%.

## 3. Results

### 3.1. Real-time PCR efficiency

The standard curve constructed from the base 10 serial dilution of the ACH-2 cell, for the IN and LTR targets of HIV-1 and the internal control of the CCR5 gene, allowed the determination of the efficiency of the assay, the linearity and the protocol of collaboration (R^2^). The assay efficiency for each target tested is shown in Table 3.

**Table 3:** Efficiency of qPCR (IN, LTR, and CCR5), slope and correlation coefficients (R^2^) to detect HIV-1 proviral DNA.

|  | IN | LTR | CCR5 |
| --- | --- | --- | --- |
| Efficiency % | 99 | 107 | 97 |
| Slope | -3.348 | -3.172 | -3.358 |
| Linearity-$R^2$ | 0.998 | 0.999 | 0.998 |

The graphic analyzes linear regression plots and amplification plots are shown in Figure 2.

**Figure 2.**
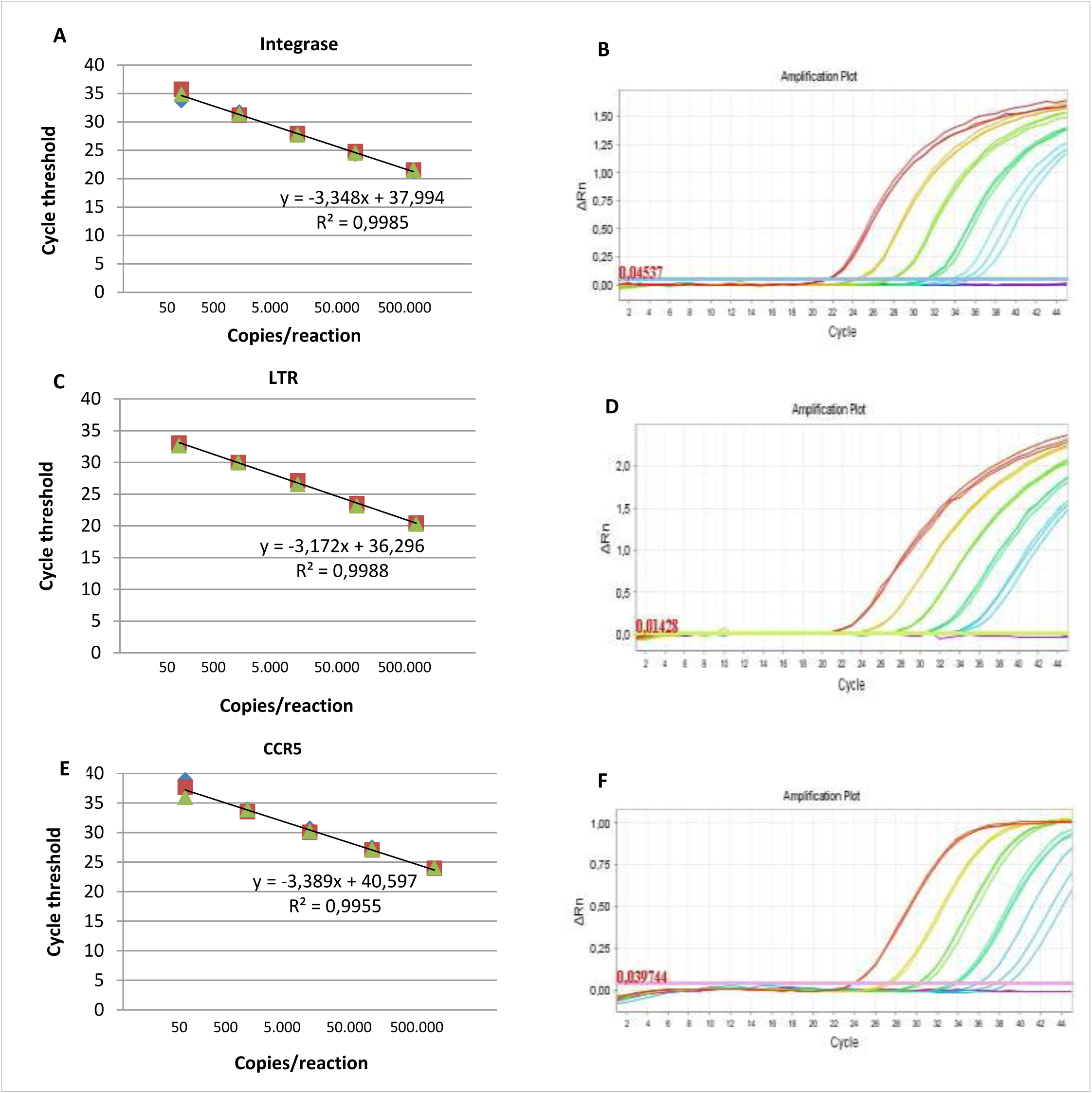
Standard curve containing five-point dilution target IN, LTR, and CCR5 (endogenous internal control) (A, C, and E), respectively. The assay was performed in triplicate. Blue diamond, 1st replica; Red square, 2nd repetition; Green triangle, 3rd replica. Amplification graph containing the five dilution points of the ACH-2 cell (B, D and F), respectively. Each color represents a concentration/copy number per reaction: red 5×10^5^, yellow 5×10^4^, light green 5×10^3^, dark green 5×10^2^, blue 5×10^1^.

### 3.2. Precision and inter-assay reproduction

Assay results obtained by two different analysts showed consistency in qPCR reproduction for the detection of HIV-1 IN (R^2^ 0.95) and LTR (R^2^ 0.96) targets.

Table 4 shows inter-assay reproducibility for detecting HIV-1 proviral DNA (IN and LTR). DNA samples of different analyte concentrations were tested in duplicate and different experiments, for five consecutive days. The results show little variation and a good correlation between positivity and detection.

**Table 4:**
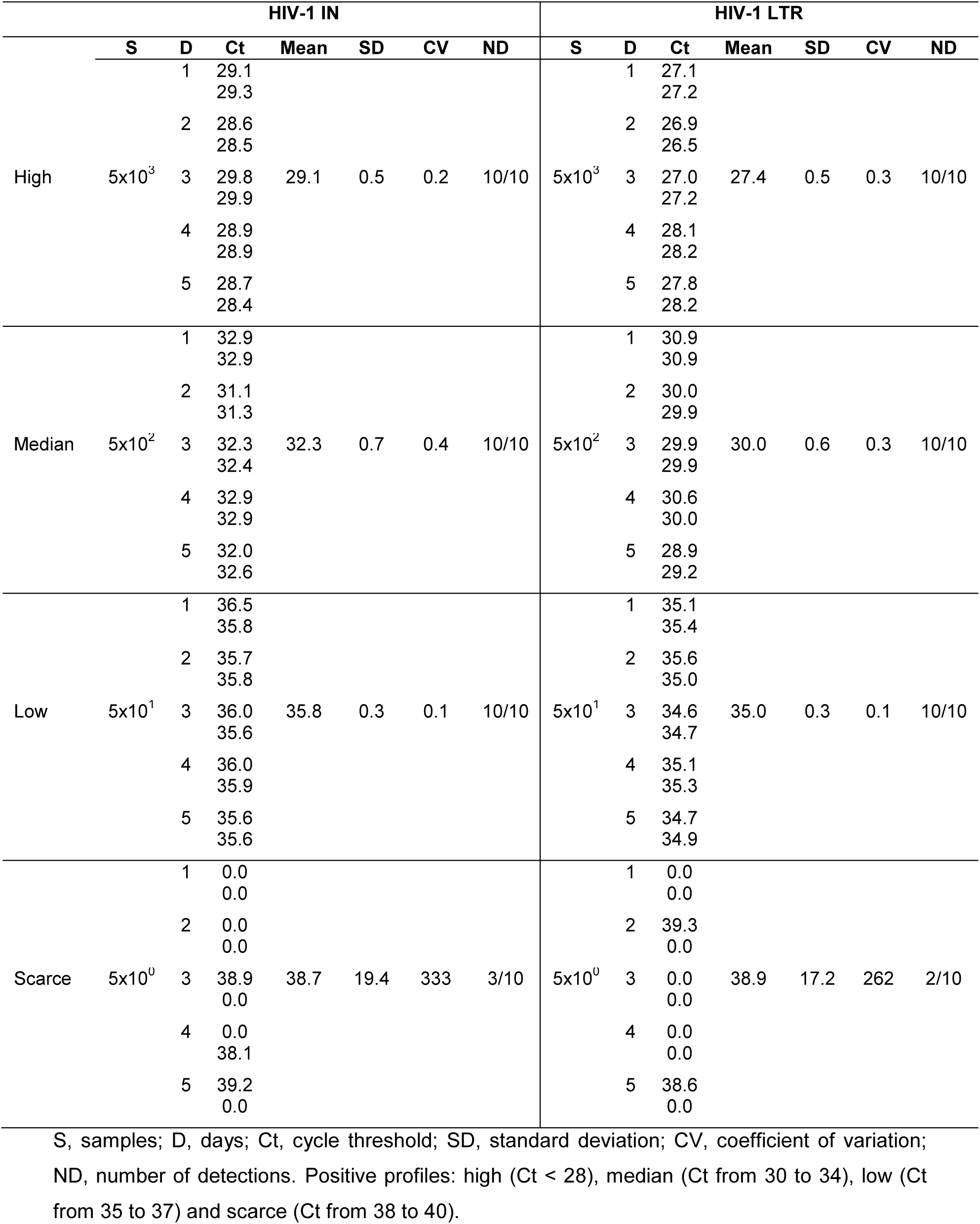
Inter-assay reproducibility (IN and LTR) for HIV-1 proviral DNA detection using DNA samples with different concentrations of the analyte, in duplicate and on five consecutive days of analysis.

### 3.3. Limit of Detection

The LOD was established at a concentration of 40 copies/reaction, as it was the lowest detectable concentration in 95% of the replicates tested (Table 5).

**Table 5:** Determination of detection limits (LOD) for IN and LTR targets.

| Copies/reaction | Integrase |  |  |  |  | LTR |  |  |  |  |
| --- | --- | --- | --- | --- | --- | --- | --- | --- | --- | --- |
|  | Ct mean | SD | CV | ND | % | Ct mean | SD | CV | ND | % |
| 50 | 36.0 | 0.4 | 0.1 | 20/20 | 100 | 34.6 | 0.5 | 0.3 | 20/20 | 100 |
| 40 | 36.9 | 0.4 | 0.1 | 19/20 | 95 | 36.1 | 0.5 | 0.2 | 19/20 | 95 |
| 30 | 37.3 | 0.4 | 0.2 | 18/20 | 90 | 36.0 | 0.5 | 0.3 | 17/20 | 85 |
| 10 | 38.0 | 0.3 | 0.1 | 13/20 | 65 | 37.4 | 0.7 | 0.4 | 12/20 | 60 |
| 5 | - | - | - | 0/20 | 0 | - | - | - | 0/20 | 0 |

SD, standard deviation; CV, coefficient of variation; ND, number of detections; % Detection percentage.

### 3.4. Evaluation of real-time PCR performance on clinical samples

The group of adult HIV-1 infected patients (aviremic, low viremia, and high viremia) was composed of 52/72 (72%) males, median age 38 years (IQR 30 – 47 years), median CD4+ T cells of 491 cells/mm^3^ (IQR 284 - 748 cells/mm^3^), median time of diagnosis of 3 years (IQR 29 days - 9 years). 66/72 (92%) of the patients were undergoing treatment, median treatment time of 3 years (IQR 2 months – 8 years), with the most frequent therapeutic regimens: tenofovir/lamivudine/dolutegravir (TDF/3TC/DTG) (35%) and tenofovir/lamivudine/efavirenz (TDF/3TC/EFV) (17%). Proviral DNA was detected in 86%, 82%, and 87% of adult patients with VL TND or <LLOQ, VL < 1000 copies/mL and VL > 1000 copies/mL, respectively. The results obtained in qPCR are presented in Table 6.

**Table 6.** Performance of qPCR in adult patients infected with HIV-1, divided by group according to VL.

| qPCR | VL TND or <LLOQ | VL < 1000 copies/mL | VL > 1000 copies/mL | Total |
| --- | --- | --- | --- | --- |
| Detected | 19 | 22 | 20 | 61 |
| No detected | 3 | 5 | 3 | 11 |
| Total | 22 | 27 | 23 | 72 |
| Detection % | 86.4 | 81.5 | 86.9 | 85.3 |

TND, target not detected; <LLOQ, lower limit of detection

Regarding the group of children infected with HIV-1 through vertical transmission included in the study (n=30), 21 (67%) were female, the median age was 4 years (IQR 10 months - 6 years), 3 / 30 patients were aviremic at the time of collection (VL TND or <LLOQ) 5/30 patients had VL < 1000 copies/mL, 22/30 patients had VL > 1000 copies/mL, the median TCD4+ cell count was of 1214 cells/mm^3^ (IQR 740 - 2386 cells/mm^3^), median infection time of 1 year (IQR 1 month – 5 years). 27/30 (90%) were undergoing treatment, median treatment time of 1 year (IQR 4 months – 7 years), with the most frequent therapeutic regimens being: zidovudine/lamivudine/lopinavir/ritonavir (AZT/3TC/LPV/RTV) (30%) and zidovudine/tenofovir/raltegravir (AZT/TDF/RAL) (13%). Of the 30 HIV-1 infected pediatric patient samples analyzed, proviral DNA was detected in 26/30 (86.6%) samples.

Clinical sensitivity and specificity were determined with 102 samples from HIV-1 infected patients and 23 samples from uninfected individuals. Proviral DNA was detected in 87/102 samples from HIV-1 positive patients, in both or only one of the targets (IN and LTR).

#### Proviral DNA in any of the HIV-1 negative patient samples analyzed

The assay evaluated presented a diagnostic sensitivity of 85.3% and specificity of 100%. Considering a disease prevalence of 1% (higher than the Brazilian national prevalence), the assay has an accuracy of 99.8%, a positive predictive power of 100%, and a negative predictive power of 99.8% (Table 7). In environments where HIV prevalence is 20%, the accuracy of the assay is 97.1% and the negative predictive power is 96.4%.

**Table 7.** Sensitivity and specificity of qPCR, evaluated using samples from HIV-1 infected and non-infected individuals.

| Patient HIV infection status | qPCR |  |  | Sensitivity (%) | Specificity (%) |
| --- | --- | --- | --- | --- | --- |
|  | Positive | Negative | Total |  |  |
| Infected | 87 | 15 | 102 | 87.2 (79.7-92.6) | 100 (85.2-100) |
| No infected | 0 | 23 | 23 |  |  |
| Total | 87 | 38 | 125 |  |  |

Calculations computed using https://www.medcalc.org/calc/diagnostic_test.php. Values in parentheses are the 95% confidence intervals.

In 15/102 samples from patients infected with HIV-1, there was no detection of proviral DNA in any of the IN and LTR targets (false-negative). 12/15 had a low concentration of total DNA (<20ng/µL), a condition that can directly interfere with the performance of the assay in detecting the target (Table 8).

**Table 8:** Clinical and laboratory data, total DNA concentration, 260/280 ratio and Ct CCR5 internal control of cases of patients infected with HIV-1, but HIV-1 proviral DNA was not detected by qPCR (false-negative).

| ID | Age (months) | VL<br>(copies/mL) | TCD4<br>(cell/mm<sup>3</sup>) | ARV | DNA ng/μL | 260/280 | Copies<br>CCR5 |
| --- | --- | --- | --- | --- | --- | --- | --- |
| 47122<sup>4</sup> | 2 | 51 | 2640 | AZT/3TC/LPVr | 2.0 | 2.4 | 22.6 |
| 48221<sup>3</sup> | 308 | 6609 | 653 | No treatment | 2.3 | 2.7 | 30.8 |
| 10022<sup>2</sup> | 575 | 71 | 205 | ATV/TDF/3TC/RTV | 2.8 | 2.0 | 34.1 |
| 12722<sup>2</sup> | 546 | 84 | 699 | ATV/TDF/3TC/RTV | 3.7 | 1.9 | 31.2 |
| 44021<sup>3</sup> | 312 | 155313 | 8 | No treatment | 4.3 | 3.1 | 28.2 |
| 1320<sup>1</sup> | 467 | TND | 558 | TDF/3TC/DTG | 5.8 | 2.6 | 27.8 |
| 13822<sup>2</sup> | 841 | 158 | 604 | TDF/3TC/EFV | 6.9 | 1.5 | 27.8 |
| 53521<sup>4</sup> | 29 | 8703 | 1510 | AZT/TDF/RAL | 9.3 | 2.4 | 28.7 |
| 26821<sup>2</sup> | 527 | 100 | 568 | TDF/3TC/EFV | 12.3 | 3.8 | 25.9 |
| 6021<sup>4</sup> | 22 | 31566 | 1261 | AZT/3TC/NVP | 17.3 | 2.7 | 24.8 |
| 4022<sup>2</sup> | 670 | 71 | 389 | TDF/3TC/EFV | 18.1 | 1.8 | 26.3 |
| 44022<sup>1</sup> | 525 | TND | 439 | TDF/3TC/DTG | 18.9 | 1.6 | 25.9 |
| 32821<sup>3</sup> | 258 | 4935 | 389 | No treatment | 32.2 | 1.9 | 25.5 |
| 2420<sup>4</sup> | 9 | <LLOQ | 3725 | AZT/3TC/NVP | 32.2 | 2.3 | 24.8 |
| 2721<sup>1</sup> | 294 | TND | 619 | TDF/3TC/EFV | 43.6 | 2.1 | 24.1 |

TND, Target not detected; <LLOQ- Lower Limit of Detection; Lymphocyte TCD4 count (cell/mm<sup>3</sup>); ARV, Antiretroviral Therapy; Ct, cycle threshold; AZT, zidovudine; 3TC, lamivudine; LPVr, lopinavir boosted with ritonavir; ATV, atazanavir; TDF, tenofovir; DTG, dolutegravir; RAL, raltegravir EFV, efavirenz NVP, nevirapine.
<sup>1</sup>Patient Sample VL TND or <LLOQ; <sup>2</sup>Sample patients with low viremia; <sup>3</sup>Patient sample with high viremia; <sup>4</sup>Sample of HIV-1 infected pediatric patient.

### 3.5. Detection of different HIV-1 subtypes

Of the 102 samples included in the study, 48/102 samples had amplification of the IN target by nested PCR, and in 44/48 genetic sequences of HIV-1 IN region were obtained. In addition to these sequences. another 14 samples not sequenced from the proviral DNA had a genetic sequence obtained from plasma (amplification of viral RNA) totaling 58 HIV-1 genetic sequences (44 - proviral DNA and 14 - viral RNA). Viral sequences were edited in Recall (http://pssm.cfenet.ubc.ca), aligned, and analyzed in Bio Edit v. 7.0.

Sequences were analyzed for subtype using the REGA HIV-1 Subtyping Tool (Version 3.0). Most sequenced samples were of subtype B (69%), followed by subtype C (15.5%), F (8.6%), BF (3.4%), AE (1.7%) and G (1.7%), concordant with the prevalence of subtypes in the state of São Paulo, Brazil (Arruda et al., 2018). When evaluating the qPCR detection rate according to the HIV-1 subtype, we observed that qPCR was able to detect a variety of HIV-1 subtypes and CRFs (Table 9).

**Table 9.** Comparison between Ct values obtained in single-step qPCR assays and Nested qPCR for different concentrations of copies/reaction, Ct values obtained in triplicates, mean, standard deviation and variance.

| Copies/reaction | qpCR |  |  |  |  |  | Nested qPCR |  |  |  |  |  |
| --- | --- | --- | --- | --- | --- | --- | --- | --- | --- | --- | --- | --- |
|  | 1. | 2. | 3. | Mean | DP | VAR | 1. | 2. | 3. | Mean | DP | VAR |
| 5 x 10<sup>5</sup> | 21,3 | 21,5 | 21,5 | 21,4 | 0,1 | 0,01 | 13,1 | 12,9 | 13,2 | 13,0 | 0,1 | 0,02 |
| 5 x 10<sup>4</sup> | 24,3 | 24,7 | 24,5 | 24,5 | 0,2 | 0,03 | 16,0 | 16,1 | 16,2 | 16,1 | 0,1 | 0,01 |
| 5 x 10<sup>3</sup> | 27,6 | 27,9 | 27,7 | 27,7 | 0,2 | 0,02 | 19,5 | 19,6 | 19,6 | 19,6 | 0,1 | 0,00 |
| 5 x 10<sup>2</sup> | 31,7 | 31,1 | 31,3 | 31,4 | 0,3 | 0,06 | 23,7 | 23,8 | 23,7 | 23,7 | 0,1 | 0,00 |
| 5 x 10<sup>1</sup> | 33,7 | 35,7 | 34,8 | 34,7 | 0,3 | 0,67 | 26,9 | 26,8 | 26,5 | 26,7 | 0,1 | 0,03 |

The tests were carried out in triplicate (Ct 1,2 and 3), SD – Standard deviation, VAR – variance.

### 3.6. Nested qPCR standard curve

The standard curve constructed for the HIV-1 IN target in Nested qPCR was performed to analyze the slope, R², and efficiency parameters. Nested qPCR for the integrase target achieved an efficiency of 93%, slope of −3,500, and R^2^ 0.9974. Figure 3.

**Figure 3.**
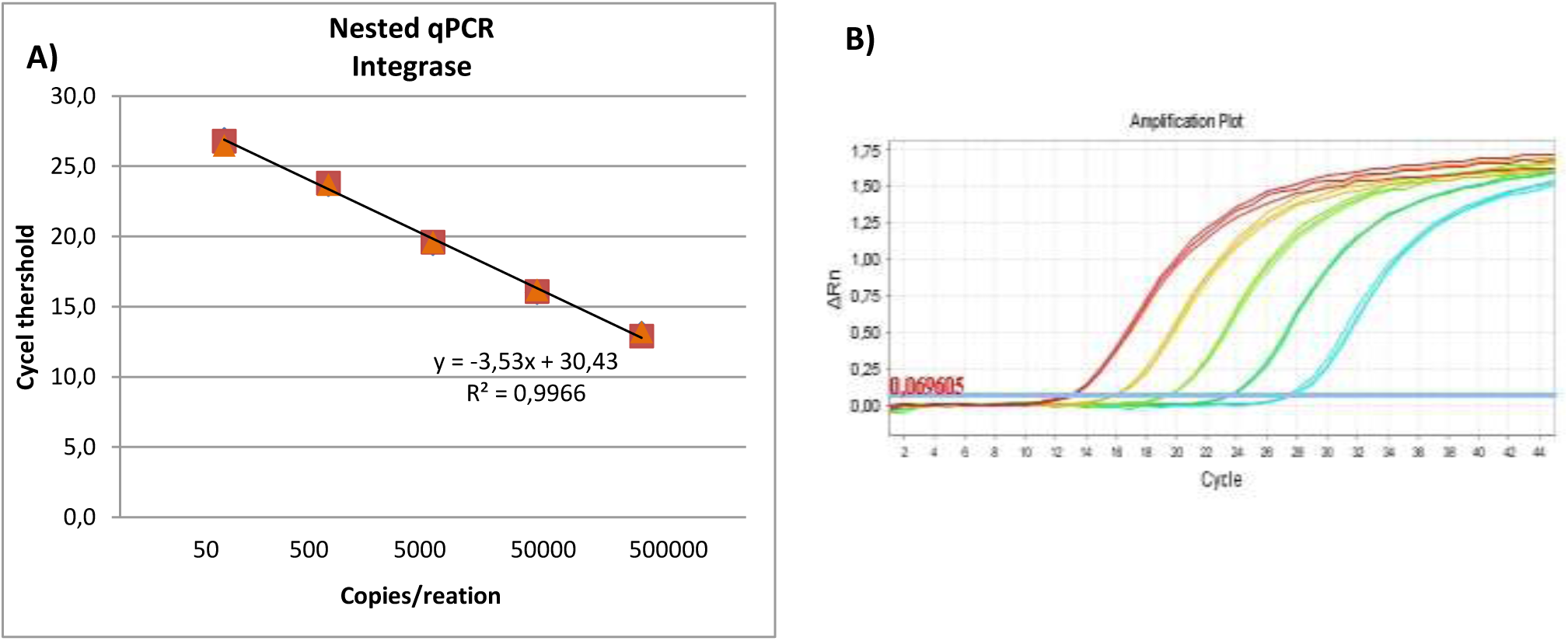
A) Standard curve graph containing five points of the Nested qPCR dilution, integrase target HIV-1. The assay was performed in triplicate. Blue diamond – 1st replicate; Red square – 2^nd^ replicate; Orange triangle – 3rd replicate. B) Amplification graph of the HIV-1 integrase target containing the five ACH-2 cell dilution points. Each color represents a concentration: red 5×10^5^, yellow 5×10^4^, light green 5×10^3^, dark green 5×10^2^, blue 5×10^1^.

### 3.7. Analytical sensitivity: Limit of detection (LOD) of Nested qPCR

The determination of the LOD of the Nested qPCR was carried out using concentrations of 50, 40, 30, 10, and 5 copies of proviral DNA - ACH-2 cell, the assays were carried out in 20 replicates. The detection limit was established at a concentration of 30 copies/reaction, as this was the lowest concentration detected in the assay in 95% of the replicates tested.

### 3.8. Performance evaluation of Nested qPCR

Serial dilutions of the ACH-2 cell were used to compare qPCR (single step) and Nested qPCR (two steps), targeting the HIV-1 IN region. All assays were performed in triplicate (Table 10).

**Table 10.**
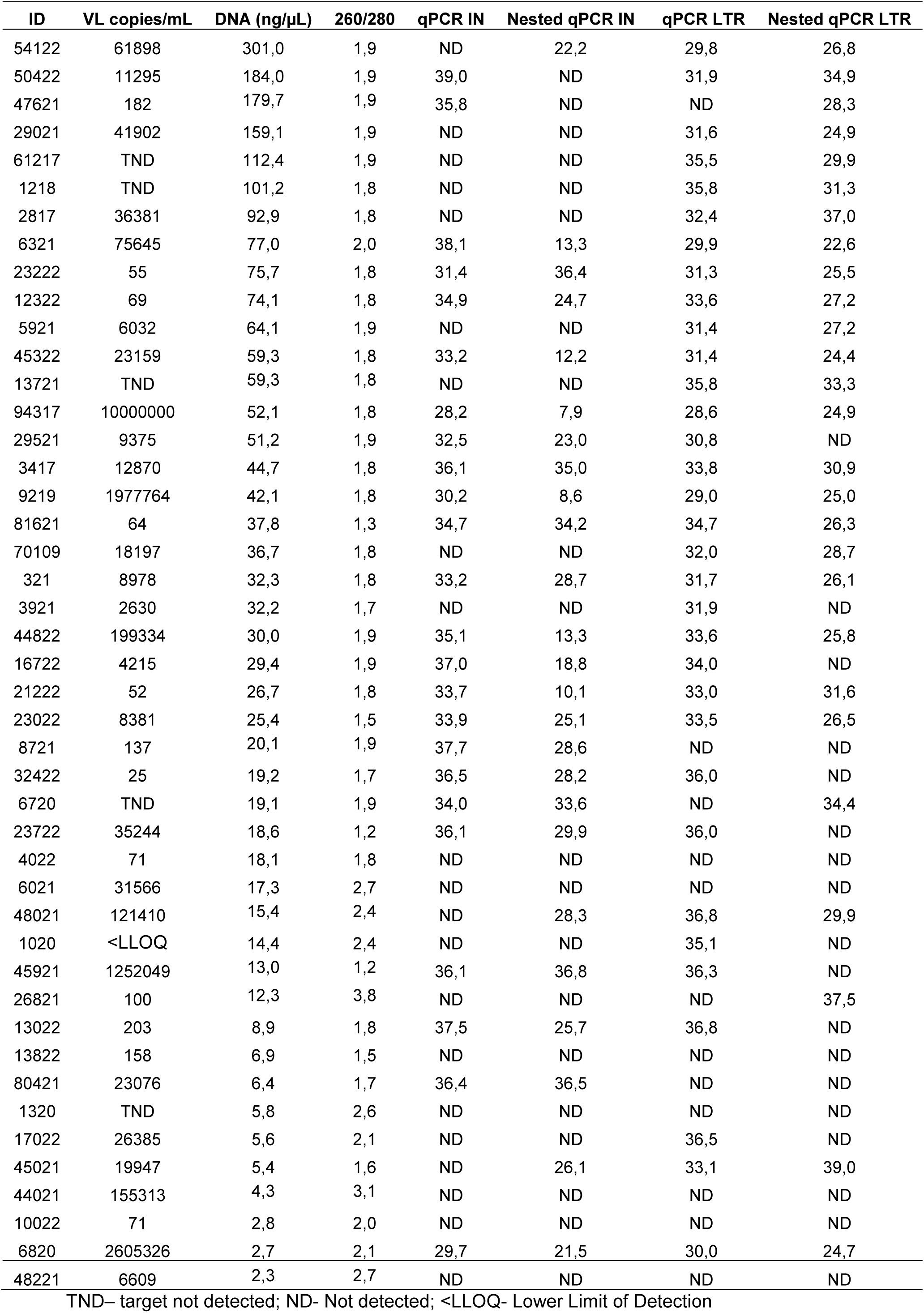
Samples from HIV-1 infected patients analyzed by qPCR and Nested qPCR.

Comparisons between linear regression plots of single-step qPCR and Nested qPCR (Figure 4) demonstrate that pre-amplification did not interfere with the linearity of the standard curves across the entire range of serial dilutions (R^2^ > 0.99).

**Figure 4.**
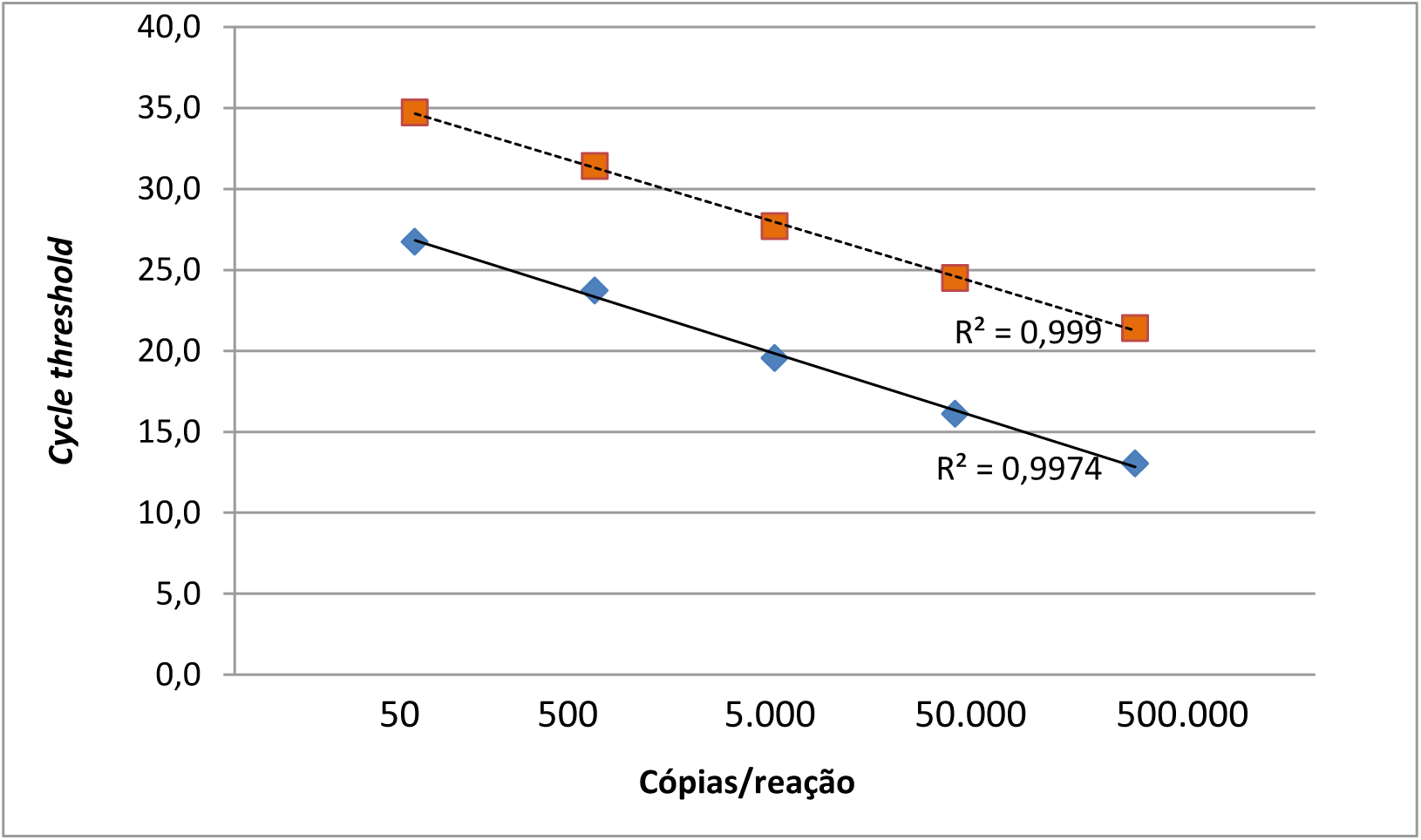
Comparison of qPCR and Nested qPCR standard curves. The tests were carried out in triplicate, the graph was constructed based on the average Ct values of the replicates. Dotted line and orange squares qPCR (single step), solid line and blue diamonds Nested qPCR (two steps).

### 3.9. Detection of HIV-1 proviral DNA in clinical samples - Nested qPCR

44 clinical samples from HIV-1 infected patients analyzed by qPCR (single step) were selected to also be analyzed by Nested qPCR (two steps). Assays were performed in duplicate, targeting the Integrase and LTR regions of HIV-1. 15/44 samples were from pediatric patients, 13/44 patients with high viremia, 11/44 patients with low viremia, 5/44 patients with VL TND or < LLOQ. Three samples from non-HIV-1 patients were included and used as a negative control.

As we can see, there was a sudden drop in the Ct value obtained when comparing the results of the two assays, qPCR, and Nested qPCR (Table 11).

In most cases, a significant drop in the Ct value was observed when comparing the two assays, increasing the probability of detection in cases where proviral DNA is scarce. In three samples analyzed from HIV-1 infected patients in which the IN target was not detected by qPCR, they were detectable by Nested qPCR (ID 54122; 48021; 45021), and the same occurred with the LTR target (ID 47621; 6720; 26821).

No amplification of HIV-specific signals was observed in HIV-1 negative samples, with CCR5 positive in 100%.

## 4. Discussion

We describe a comprehensive, simple, highly sensitive, specific, and reproducible assay for the detection of HIV-1 proviral DNA from buffy coat cells obtained from routine whole blood samples leftovers.

One of the advantages of the assay is that it is carried out using mononuclear blood cells collected from the buffy coat, without the need for Ficoll-Hypaque to separate PBMC (Désiré et al., 2001; Zhao et al., 2002; Lillo et al., 2004). PBMC separation tests were carried out using Ficoll-Hypaque in some samples (data not shown) in an attempt to improve the DNA concentration obtained in the extraction, however, the results of the total DNA dosage and qPCR were not superior when compared to the PBMC collection performed directly from the buffy coat, in addition to being a time-consuming technique that requires cell separation steps that may not be provided by routine laboratory centrifuges and representing an additional expense for the assay.

The evaluated qPCR showed satisfactory performance for all tested targets, a wide dynamic range, and a detection limit of 40 copies/reaction of HIV-1 proviral DNA. Detection of a low copy number of HIV-1 proviral DNA makes the method potentially useful for use in recent infections and infants with extremely low levels of proviral DNA due to early treatment or antiretroviral prophylaxis to prevent mother-to-child transmission. However, the actual performance of the test in these situations was not properly evaluated in this study.

One of the main characteristics of HIV-1 is its extreme genetic variability attributable to high rates of mutation, subtype-related polymorphisms, and frequent recombination (Rouet et al., 2007; Santoro et al., 2013). Before starting the qPCR tests, the primers and probes used in the study were analyzed for their specificity and ability to pair with HIV-1 proviral DNA sequences, analyzes were performed using sequences (total of 410) from all subtypes of various countries, available in the Los Alamos database. All oligonucleotides used showed excellent agreement in sequence conservation in the integrase and LTR region. As noted by Cillo et al. (2014) there is excellent agreement in the conservation of the sequence in the integrase region except the substitution of a nucleotide present in the binding site of the ISCA forward primer. However, this condition did not prove to be a limitation for HIV-1 detection when using the primer.

The use of two conserved regions of the viral genome (IN and LTR) favored the increase in the sensitivity of the assay, helping to overcome limitations caused by the genetic diversity of HIV-1, as observed in the assay developed by Avettand-Fènoël and collaborators, 2009.

Sensitivity and specificity are of paramount importance for qualitative assays designed for diagnostic purposes, our qPCR showed a sensitivity of 85.3% and a specificity of 100% estimates that were tested using a range of patient characteristics and plasma viremia levels. qPCR was able to detect HIV-1 proviral DNA in HIV-1 infected adult patients with undetectable plasma VL or below the minimum detection limit (<20 copies/mL). The detection of HIV-1 proviral DNA in samples from ARV-suppressed patients is highly challenging due to the low abundance of HIV-1 DNA and viral heterogeneity in these patients (Avettand-Fènoël et al., 2016). The detection among cases with TND or below the limit of detection at similar rates of viremic patients reassures the usefulness of the test when RNA detection cannot provide HIV diagnosis confirmation. The impact of a long time of viral suppression in these detection rates needs however further evaluation. Moreover, the detection and quantitation of HIV-1 viral reservoir in proviral DNA are of paramount importance in HIV-1 cure research (Mellberg et al., 2017). Although the present study focuses on the qualitative detection of HIV-1 proviral DNA, the assay can be adapted for quantitative measurements of proviral DNA in whole blood. The assay may thus provide semi-quantitative information on Proviral frequency among PBMCs, but this needs further analysis.

Most cases of false-negative results observed in the study can be explained by the low concentration of total DNA present in the sample, such a condition can directly interfere with the assay’s ability to detect the target, following the Poisson distribution.

In addition, another aspect that must be considered for the non-detection of proviral DNA is related to the low levels of the target, such levels would be below the detection limit of the assay. Virological and/or host factors, as well as initiation of an antiretroviral regimen, may affect the detection of HIV-1 proviral DNA. Very low levels of HIV-1 proviral DNA have been seen in slow progressors and elite controllers: patients who have sustained natural control of HIV replication and no disease progression after years of infection without taking ARVs (Lambotte et al., 2005; Balasubramanian et al., 2017).

Methods for detecting proviral DNA need to be sensitive, but also subtype-independent, as many research and clinical settings are dealing with a diversity of HIV-1 subtypes (Mellberg et al., 2017). The evaluated assay detected a variety of HIV-1 subtypes and CRFs, including B, C, F, BF, and AE, indicating that the assay is robust enough to detect most if not all HIV-1 group M variants, however, the analysis of a larger number of samples of different subtypes is necessary.

As seen in the studies by Pasternak et al., 2008 and Kibirige et al., 2022, which combined the accuracy and precision of qPCR with the sensitivity of Nested PCR, managed to develop an assay with superior sensitivity when compared to single-step qPCR. We based this protocol on these previous studies and were able to develop an assay like Nested qPCR, with the use of an additional amplification step to increase the sensitivity of the assay, 30 copies/reaction of HIV-1 proviral DNA can be detected by Nested qPCR compared to the detection limit of 40 copies per single-step qPCR reaction. In addition to the significant drop in the Ct value observed in most cases when comparing the two assays, three samples in which proviral DNA was not detected in qPCR were detected by Nested qPCR, showing that the use of an additional amplification step allows enrichment of the target increasing the chances of detection.

As qPCR allows quick and easy processing of a large number of samples, assays for the detection of HIV-1 proviral DNA can be used as a complete tool, being an alternative to commercially available assays. Currently in Brazil, there is only one commercial assay for the detection of available HIV-1 proviral DNA, the GeneXpert HIV-1 Qual, the assay achieves excellent sensitivity and specificity, 94.1% and 99.8%, respectively (Opollo et al., 2018), however, it requires specific equipment, and kits are expensive. We carry out the calculations briefly to find out the costs per qPCR test, each test costs approximately seven us dollars, including the DNA extraction stage. With commercial kits, each test would cost approximately 37 US$. Thus, the In-house qPCR has a cost of 5 times lower.

The assays evaluated in this study had a low limit of detection, suggesting that they are appropriate for the early diagnosis of HIV-1 infection in infants. Good assay performance was also demonstrated in HIV-1-infected adults. Additionally, the assay may be useful in determining the status of HIV-1 infection when serology results are indeterminate, especially useful to allow patients to continue therapy or prophylaxis.

## 5. Conclusion

The evaluated qPCR showed satisfactory performance for detecting HIV-1 proviral DNA, proving to be an appropriate complementary tool for early pediatric diagnosis, diagnostic elucidation, as well as monitoring in cases where HIV-1 RNA is not detectable in plasma.

## Data Availability

All data produced in the present work are contained in the manuscript

